# PREVALENCE OF ABNORMAL SEMINAL FLUID AND ASSOCIATED FACTORS AMONG PATIENTS ATTENDING FERTILITY CLINICS IN OSOGBO: A CROSS SECTIONAL STUDY

**DOI:** 10.1101/2024.08.01.24311347

**Authors:** Kehinde Awodele, Sunday Charles Adeyemo, Eniola Dorcas Olabode, Adeniyi Olaonipekun Fasanu, Lanre Olaitan, Akintunde Rasaq Akindele, Olufunso Abidemi Olagunju, Adeola Dorcas Aderinwale

**Affiliations:** Obstetrics and Gynecology Department, Uniosun Teaching Hospital, Osogbo, Nigeria; Institut Superieur de Sante, Niamey, Niger Republic; Department of Community Health, Obafemi Awolowo University, Ile-Ife, Nigeria; Department of Obstetrics and Gynecology, Osun State University, Osogbo, Nigeria; Department of public health/fb. University of Ilorin; Department of Pediatrics, Osun State University, Osogbo, Osun State, Nigeria; Primary Health Care Unit, Adeleke University, Ede, Nigeria

**Keywords:** Male Infertility, Implications, Seminal fluid Analysis, Sperm count

## Abstract

**Background:** Infertility is a global health issue that affects millions of couples worldwide, In Nigeria, the prevalence of infertility is particularly high underscoring the need for a better understanding of the factors contributing to male infertility at which Seminal Fluid Analysis (SFA) is the most important characteristics. This study aimed to assess the prevalence of abnormal seminal fluid and its associated factors among patients attending fertility clinics in Osogbo, Nigeria.

**Methods and Findings:** A descriptive cross-sectional study design was used among male patients attending fertility centers in Osogbo, Osun State. Fisher’s formula (n=z^2^pq/d^2^) was used to determine the sample size. 305 respondents were selected using multistage sampling technique. Pre-tested questionnaire was used to collect data from respondents. Patients who consented to the study were also made to undergo seminal fluid analysis. The results were analysed using IBM Statistical Product for Service Solution (SPSS) version 27. Descriptive statistics was used for all variables. Bivariate and multivariate analysis were done at p<0.05 as level of significance. Majority of the respondents, 257 (84.3%) had at least one abnormality of Seminal fluid. Multiple regression analysis revealed that respondents who were habitual drunkard were about five times more likely to have at least one abnormality in their seminal fluid (OR: 4.990, p: 0.004, C.I.: 1.688-14.749) and smokers were three times more likely to have at least one abnormality in their seminal fluid (p=0.005, OR=3.300 and C.I=1.396-4.273). Also, respondents with history of sexually transmitted infection were 3.5 times more likely to have at least one abnormality in their seminal fluid (p=0.039, OR=3.595 and C.I=1.072-14.146).

**Conclusions:** The study observed high prevalence of abnormal seminal fluid which was significantly associated with lifestyle habits such as smoking, alcohol as well as previous history of sexually transmitted infection. The study recommended that advocacy program for healthy lifestyle, early screening and public health education will further reduce the burden of abnormal seminal fluid and by implication, infertility among couples.

## INTRODUCTION

Infertility is a global health issue that affects millions of couples worldwide. The inability to conceive a child after a year of regular, unprotected intercourse is a distressing experience for many couples, leading to emotional, psychological, and social challenges. [1] In Nigeria, the prevalence of infertility is particularly high, with studies showing that around 20-30% of couples face fertility challenges. [2]

One of the most important aspects of male infertility is the quality and characteristics of the seminal fluid. Seminal fluid, also known as semen, is a complex mixture of spermatozoa and secretions from the male reproductive glands, including the seminal vesicles, prostate, and bulbourethral glands. Seminal fluid plays a crucial role in the fertilisation process, providing a supportive environment for sperm transport, nourishment, and protection against the female reproductive tract’s hostile conditions. [3]

Infertility is a significant public health concern affecting millions of couples worldwide, with male infertility accounting for approximately half of these cases.[4] In Southwest Nigeria, particularly, there is a noticeable gap in research regarding the seminal fluid analysis (SFA) and its implications on male infertility among patients attending fertility clinics [5] Therefore, this study aims to assess the prevalence of abnormal seminal fluid among male patients attending fertility centers in Osogbo, Osun State, Nigeria.

## METHODS

A descriptive cross-sectional study design was used in the study. The study population were male patients attending fertility centers in Osogbo, Osun State. Fisher’s formula (n=z^2^pq/d^2^) was used to determine the sample size. 305 respondents were selected using multistage sampling technique. Out of 10 major fertility centers in Osogbo, Osun State, 5 centers were randomly selected. Proportional allocation was used to determine the number of respondents from each center. Male patients aged 18-50 were randomly selected in each center. Pre-tested questionnaire was used to collect data from respondents. Patients who consented to the study were also made to undergo seminal fluid analysis. The semen was collected aseptically after an abstinence period of 2–7 days using sterile universal bottles in a private room near the laboratory to limit the exposure of the semen to fluctuations in temperature and to control the time between collection and analysis. The semen was then analysed in the laboratory for sperm count, motility and morphology of the semen. The data from the questionnaire were analysed using IBM Statistical Product for Service Solution (SPSS) version 27. Descriptive statistics was used for all variables. Bivariate and multivariate analysis were done at p<0.05 as level of significance.

## RESULTS

### Sociodemographic characteristics of respondents

All the respondents were married male attending the infertility clinic in Osogbo. More than half 164 (53.8%) of the respondents were with the ages of 31-40 years, while the majority, 267(87.5%) were Yoruba. 164 (53.8%) had tertiary level of education followed by secondary 83 (27.2%). About half 140 (45.9%) were skilled workers while 110 (36.1%) were unskilled.

### Prevalence of abnormal seminal fluid

Majority of the respondents, 84.3% had at least one abnormality while 15.7% have normal seminal fluid. About two-thirds 217 (71.1%) had normal sperm count (>32 million per ejaculation) while 88 (28.9%) had abnormal sperm count. Only 97 (31.8%) had normal progressive motility (>32 percent) while 208 (68.2%) had abnormal (Athenospermia) progressive motility and 178 (58.4%) were found to have abnormal morphology (Teratospermia i.e., <4%). (Figure 1)

**Figure 1:**
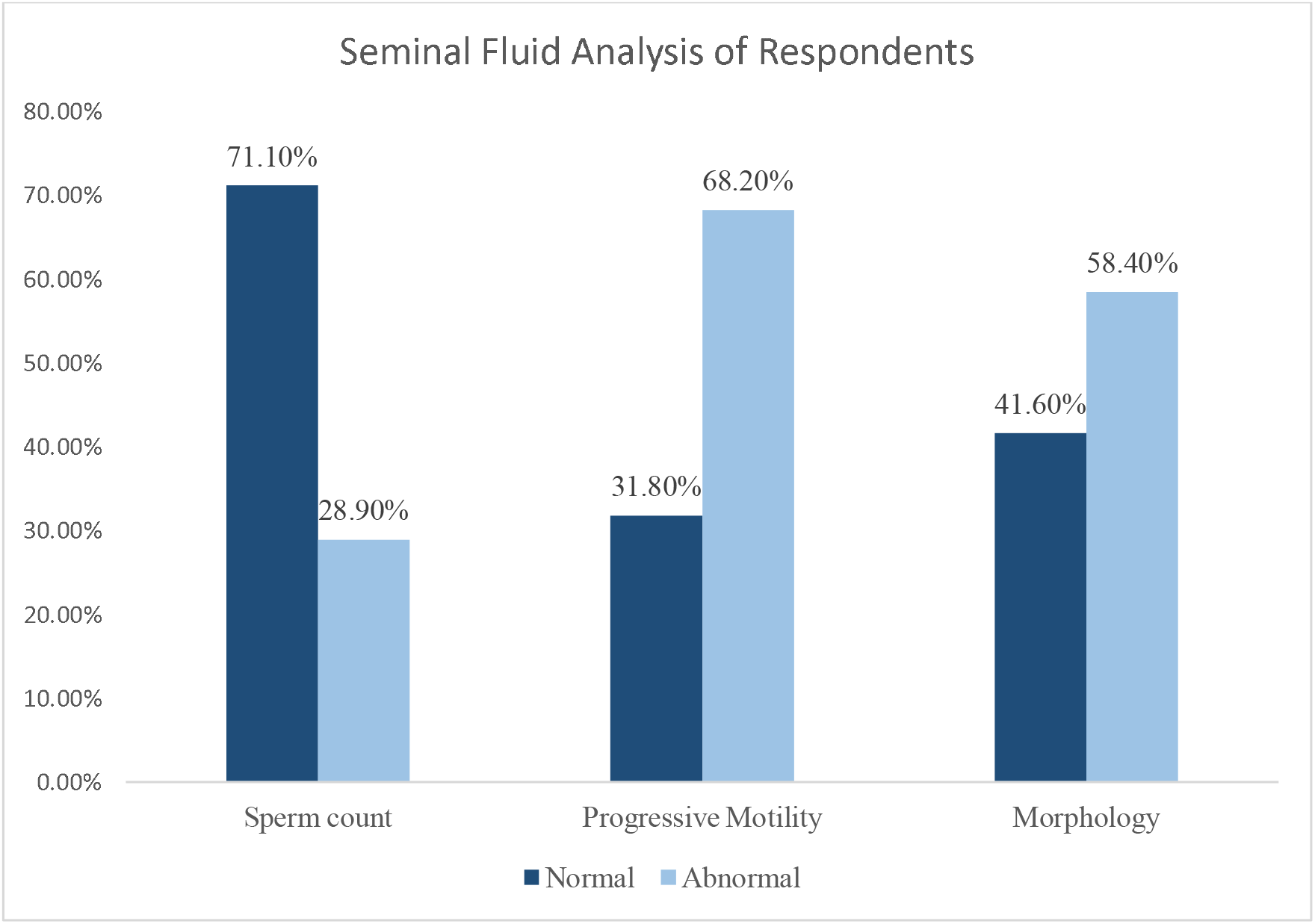
Seminal fluid Analysis of Respondents.

### Lifestyle factors of respondents

A little above one-third were known smokers (35.1), out of which 78.5% smokes cigarette and 21.5% smokes marijuana. Less than half (47.5%) were known drunkard while only 8.5% currently used drugs. The majority (68.2%) eats any type of food available while 23.6% regularly consumed carbohydrate/starch food and only 1.0% consumed vegetables. (Table 1)

**Table 1:** Lifestyle behaviours among the respondents.

| Variables | Frequency (n=305) | Percentage |
| --- | --- | --- |
| **Known smoker** |  |  |
| Yes | 107 | 35.1 |
| No | 198 | 64.9 |
| **Type of smoke used (n=107)** |  |  |
| Cigarette | 84 | 78.5 |
| Marijuana | 23 | 21.5 |
| **Known Drunkard** |  |  |
| Yes | 145 | 47.5 |
| No | 160 | 52.5 |
| **Currently using any drug** |  |  |
| Yes | 26 | 8.5 |
| No | 279 | 91.5 |
| **Type of food consumed regularly** |  |  |
| Any food | 208 | 68.2 |
| Carbohydrate/Starch | 72 | 23.6 |
| Protein | 22 | 7.2 |
| Vegetables | 3 | 1.0 |

### Association between medical history of respondents and abnormal seminal fluid among respondents

Majority of the respondents 65 (91.5%) who were diabetic had at least one abnormality in seminal fluid. Majority of the respondents 29 (96.7%) who were hypertensive had at least one abnormality in seminal fluid. Majority of the respondents 49 (90.7%) who were overweight had at least one abnormality in seminal fluid. However, there was no statistically significant association between these factors and abnormal seminal fluid. Majority of the respondents 84 (96.6%) who had history of sexually transmitted infection had at least one abnormality in seminal fluid and this was statistically significant (p<0.001). (Table 2)

**Table 2:**
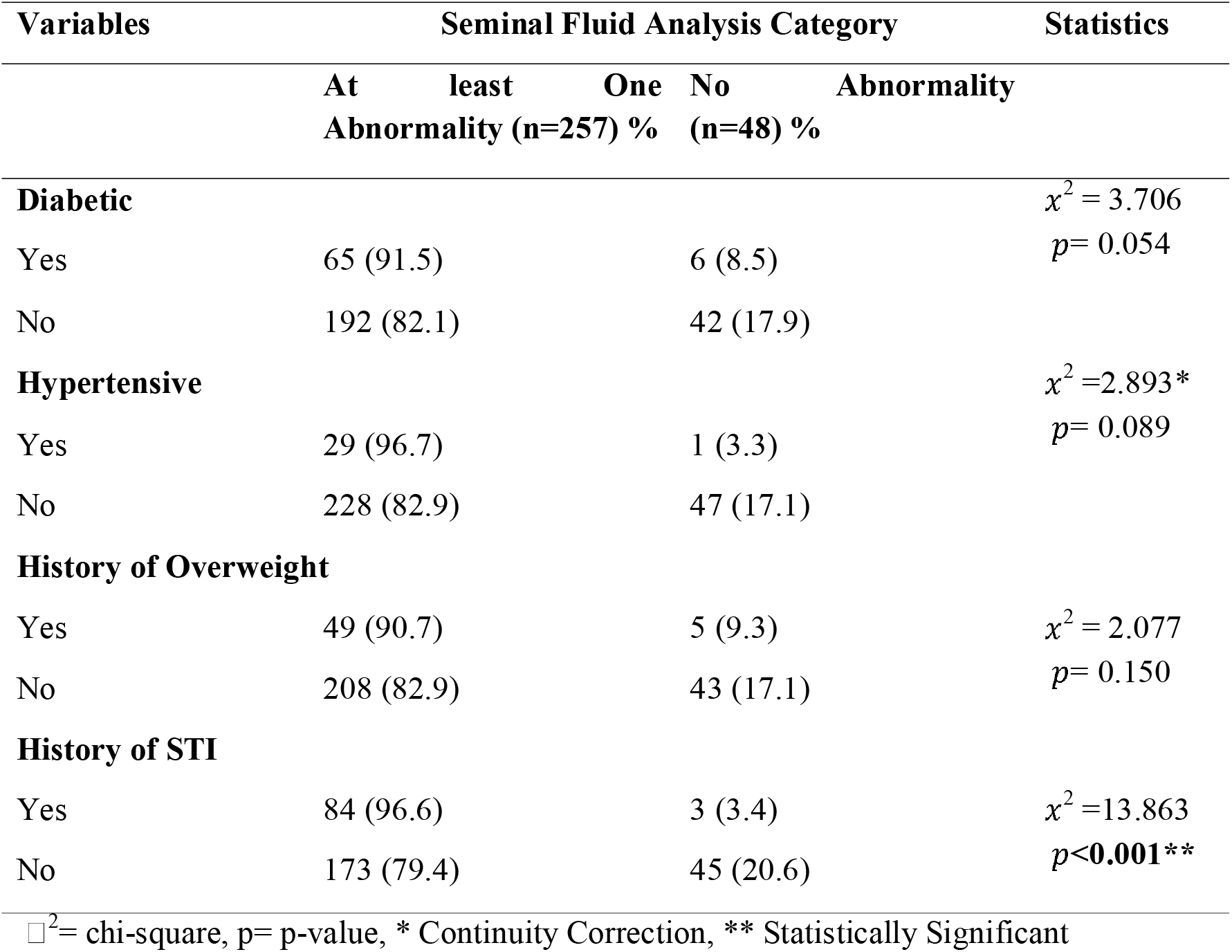
Association between medical history of respondents and abnormal seminal fluid among respondents.

### Predictors of abnormal seminal fluid among respondents

Multiple regression analysis revealed that respondents who were habitual drunkard were about five times more likely to have at least one abnormality in their seminal fluid (OR: 4.990, p: 0.004, C.I.: 1.688-14.749) and smokers were three times more likely to have at least one abnormality in their seminal fluid (p=0.005, OR=3.300 and C.I=1.396-4.273). Also, respondents with history of sexually transmitted infection were 3.5 times more likely to have at least one abnormality in their seminal fluid (p=0.039, OR=3.595 and C.I=1.072-14.146).

## Discussion

This study assessed the prevalence of abnormal seminal fluid and its associated factors among patients attending fertility clinics in Osogbo, Osun State, Nigeria. More than half of the respondents were between the ages of 31 and 40 years old while more than two-fifth were aged 21-30 years. More than half of the respondents have tertiary as their highest educational attainment. This is similar to another study conducted among male partners attending infertility clinic in a tertiary Nigerian hospital [6]

More than four-fifth of the respondents had at least one abnormality. About one-third had abnormal sperm count, which is below the WHO 2010 reference for semen analysis. [7] This proportion is also lower compared to the study of [8] where more than four-fifths were found to have abnormal sperm count. Almost 7 out of 10 respondents had abnormal (Athenospermia) progressive motility) which is more than the WHO 2010 reference of <32%. [7] Furthermore, this proportion is higher compared to another study carried out among male partners of infertile couples seeking care at the Lagos University Teaching Hospital where 55.8% had abnormal progressive motility. [9] Also, about three-fifth had abnormal morphology (Teratospermia i.e., <4%). This proportion is much higher compared to 60.8% of abnormal morphology reported in the study of [6] among male patients in a tertiary Nigerian hospital.

The predictors of abnormal seminal fluid among respondents included smoking, drinking, and previous history of sexual transmitted infection such that respondents who were habitual drunkard were about five times more likely to have at least one abnormality in their seminal fluid (OR: 4.990, p: 0.004, C.I.: 1.688-14.749) and smokers were three times more likely to had at least one abnormality in their seminal fluid (p=0.005, OR=3.300 and C.I=1.396-4.273). Also, respondents with history of sexually transmitted infection were 3.5 times more likely to have at least one abnormality in their seminal fluid (p=0.039, OR=3.595 and C.I=1.072-14.146). This was also evident in the study of [6] where age, smoking and alcohol consumption were reported as the possible risk factors for abnormal sexually transmitted infection. This suggests that substance abuse, habitual smoker, drinking habit have detrimental impact on reproductive health and fertility as it has been associated with reduced sperm count, motility, and normal morphology. It is also linked to increased levels of sperm aneuploidy, lower seminal plasma antioxidant levels, and increased oxidative damage to sperm deoxyribonucleic acid (DNA). [10]

## Conclusions

The findings indicate a prevalence of abnormal seminal fluid parameters (low sperm count, motility, and morphology) among the participants in which motility has the highest prevalence followed by morphology and sperm count respectively. Lifestyle factors such as smoking, alcohol consumption and sexually transmitted infections were found to be significantly associated with abnormal seminal fluid. The implications of abnormal seminal fluid parameters on infertile males are substantial, extending beyond physical health to encompass psychological, emotional, and social well-being. The findings from this study indicates the need for increased awareness about male infertility and accessibility to male fertility testing in order to combat the issue of infertility among couples. Further studies can assess the effect on seminal fluid analysis on male infertility.

## Data Availability

All data produced in the present study are available upon reasonable request to the authors

## Acknowledgements

The authors wish to acknowledge the respondents who gave their consent to be part of this study as well as the spouses of the authors for their understanding during the course of this study.

## Limitation

The study is limited to patients attending fertility centers in Osogbo, which can have effect on the generalization of the study. Also, due to the nature of the study as a cross-sectional study, causal relationship may not be established.

## Ethical approval and consent to participate

Ethical approvals for this study were obtained from the Ethics and Research Committee of Adeleke University, Ede, and Uniosun Teaching Hospital. The Protocol number UTH/EC/2024/06/957. All ethical principles guiding the conduct of research such as informed consent, beneficence, non-maleficence, confidentiality, justice, autonomy etc were strictly taken into consideration. All information gathered were confidential and privacy and confidentiality of the respondents were guaranteed. All respondents were made to sign the informed consent before filling the questionnaire.

## Consent for publication

Not applicable

## Availability of data and material

The data for this study is available on reasonable request from the corresponding author.

## Authors’ contribution

Kehinde Awodele-Research design, ethical compliance and data collection, Sunday Charles Adeyemo-Data Analysis and manuscripting, Eniola Dorcas Olabode- Data analysis and manuscripting, Adeniyi Olaonipekun Fasanu- Data collection and ethical compliance, Lanre Olaitan- Supervision and ethical compliance, Akintunde Rasaq Akindele- Data collection and ethical compliance, Olufunso Abidemi Olagunju- Data collection and Analysis, Adeola Dorcas Aderinwale-Data collection

## REFERENCES

[1] Zegers-Hochschild, F., Adamson, G.., De Mouzon, J., Ishihara, O., Mansour, R., and Nygren, K. (2009). The International Committee for Monitoring Assisted Reproductive Technology (ICMART) and the World Health Organization (WHO) Revised Glossary on ART Terminology,. Human Reproduction, 24, 2683–2687.

[2] Okonofua, F., Ntoimo, L., Ogungbangbe, J., Anjorin, S., Imongan, W., and Yaya, S. (2018). Predictors of women’s utilization of primary health care for skilled pregnancy care in rural Nigeria. BMC Pregnancy and Childbirth, 18(1), 1–15. 10.1186/s12884-018-1730-4

[3] Robertson, S. A., and Sharkey, D. J. (2016). Seminal fluid and fertility in women. Fertility and Sterility, 106(3), 511–519. 10.1016/j.fertnstert.2016.07.1101

[4] Agarwal, A., Mulgund, A., Hamada, A., & Chyatte, M. (2015). A unique view on male infertility around the globe. Reproductive Biology and Endocrinology, 13(1), 37.

[5] Ibitoye, B. O., Fasasi, A. O., Imosemi, I. O., Alabi, O., Olaniyan, O. T., Ibitoye, F. O., Faduola, P., Bodun, D. S., and Wike, N. Y. (2023). The spermiogram and correlation of seminal fluid parameters in patient attending fertility centre in Lagos, Southwest Nigeria. Morphologie, 107(359), 100606. 10.1016/j.morpho.2023.05.004

[6] Umar, A.., Panti, A.., Mbakwe, M., Ahmed, Y., Garba, J.., & Nnadi, D. C. (2020). The Pattern of Seminal Fluid Analysis among Male Partners Attending an Infertility Clinicin a Nigerian Tertiary Health Institution. Open Journal of Obstetrics and Gynecology, 10, 957–967.

[7] WHO. (2021a). WHO laboratory manual for the examination and processing of human semen. In WHO laboratory manual for the examination and processing of human semen (5th ed.) (5th ed.).

[8] Tilahun, T., Oljira, R., and Getahun, A. (2022). Pattern of semen analysis in male partners of infertile couples in Western Ethiopia: Retrospective cross-sectional study. SAGE Open Medicine, 10, 1–7. 10.1177/20503121221088100

[9] Makwe, C. C., Ugwu, A. O., Ojewola, R. W., and Onyeze, C. I. (2021). Seminal fluid parameters of male partners of infertile couples seeking care at the Lagos University Teaching Hospital. International Journal of Reproduction, Contraception, Obstetrics and Gynecology, 10(4), 1347. 10.18203/2320-1770.ijrcog20211109

[10] Joo, K., Kwon, Y., Myung, S., & Kim, T. (2012). The Effects of Smoking and Alcohol Intake on Sperm Quality: Light and Transmission Electron Microscopy Findings. Journal of International Medical Research, 40(6), 2327–2335. 10.1177/030006051204000631

